# Estimating the number of breakthrough COVID-19 deaths in the United States

**DOI:** 10.1101/2022.04.04.22272731

**Authors:** Jinkinson Smith

## Abstract

While there is compelling evidence of the effectiveness of COVID-19 vaccines, increasing attention has also been paid to the fact that, like all vaccines, they are not 100% effective. Therefore, some fully vaccinated people have developed “breakthrough” cases of COVID-19, and some of these individuals have died as a result. The purpose of this study was to estimate the number of fully vaccinated or “breakthrough” deaths from COVID-19 in the United States. Data was compiled from state COVID-19 dashboards and various other sources for as many states as possible. As of March 27, 2022 based on data from 46 U.S. states and the District of Columbia, an estimated minimum of 57,617 breakthrough COVID-19 deaths had occurred in the United States. Furthermore, based on this incomplete data, a total of 12.8% of all COVID-19 deaths in the included regions and time periods were among fully vaccinated individuals (whether boosted or not). Extrapolating this data to the entire United States implies that the minimum total number of such deaths as of March 27, 2022 was 79,917. Data from a MMWR article, if similarly extrapolated to the entire country, implies a significantly larger number of breakthrough deaths throughout the United States: 99,152.

## Background

The safety and effectiveness of COVID-19 vaccines is well established (e.g. Lopez Bernal et al. 2021, Zheng et al. 2022). However, breakthrough cases of COVID-19, i.e. cases of COVID-19 among individuals who had already been fully vaccinated against the disease, have also been documented extensively (e.g. Birhane et al. 2021, Lipsitch et al. 2021).

The shortcomings of the CDC’s collection of data regarding breakthrough cases of COVID-19 in the United States have been well documented (Deam & Fortis 2021, Mitropoulos 2021). These shortcomings include that not all states report data on breakthrough cases in general.

Furthermore, whether states voluntarily report data on breakthrough cases to the CDC is crucial because the CDC has not collected such data comprehensively since May 1, 2021, when it decided to only track breakthrough cases that resulted in hospitalization or death (Deam & Fortis 2021).

While previous articles have reported estimates of breakthrough case/death data in the United States, these seem to be lacking in scope and recency, so here I have attempted to comprehensively compile data on COVID-19 breakthrough infections in the United States, specifically those that resulted in death. Such data are reported with widely varying consistency and detail across individual states (and to some extent counties).

## Methods

I tried to compile data on breakthrough COVID-19 deaths from as many U.S. states (and the District of Columbia) as possible based on state COVID-19 data dashboards or other official data sources. This involved many Google searches as well as searches on the websites for individual states (e.g. ohio.gov for the state of Ohio) for terms such as “breakthrough” and “COVID-19”.

I was able to find some total number of COVID-19 breakthrough deaths for 47 United States jurisdictions (46 states and D.C). The only 4 states for which I could not find such data at least at the state level were Florida, Iowa, Kansas, and New York. Note that it was not until after I had written most of this paper that I discovered the existence of a dashboard compiled by the Rockefeller Foundation (“United States Vaccine Breakthrough Reporting Scorecard” 2022) listing the quality of publicly available reporting of COVID-19 breakthrough data by state, and assigning grades to each state in this regard. The five states I mentioned above vary in terms of their grades on the scorecard, e.g. the scorecard indicates that there is no breakthrough COVID-19 data reported from Florida or Kansas, but gives an “F” grade to Iowa while including one source for it; they also give New York and Texas a C grade and provide two links related to breakthrough data for each state. The link for Iowa (coronavirus.iowa.gov) no longer works, and the two links for New York are:

1. One for breakthrough data that only includes per capita rates of breakthrough cases and hospitalizations, and omits any data on breakthrough deaths as well as any raw numbers of breakthrough cases/hospitalizations/deaths.^2^
2. A page listing the progress of the vaccine rollout in New York with no information about breakthrough cases.^3^

Finally, the two links for Texas are:

1. One comparing per capita rates of COVID-19 cases and deaths among vaccinated versus unvaccinated people.^4^
2. A page listing the progress of the vaccine rollout in Texas with no information about breakthrough cases.^5^

After originally starting this paper I discovered a now-offline link to a document that used to be on the Texas state government website. This document lists the number of breakthrough cases and deaths for most of 2021.^6^ Therefore, I removed Texas from the list of states without breakthrough COVID-19 death data, a list that therefore was reduced to four states (as mentioned above, these are Florida, Iowa, Kansas, and New York).

For Mississippi, I was only able to look at data for (approximately) the previous month, and I had to indirectly calculate the number of breakthrough deaths and boosted deaths separately based on the total number of COVID-19 deaths that occurred during that time range (calculated from New York Times data). This is because Mississippi only reports the percentage of COVID-19 deaths during the previous month that fell into each vaccination status category: a) unvaccinated, b) fully vaccinated but not boosted and eligible for a booster, c) fully vaccinated but not boosted and not eligible for a booster, and d) fully vaccinated and boosted. A similar but distinct problem presented itself with Arkansas, which doesn’t report the total number of breakthrough deaths but does report the percentage of deaths among unvaccinated individuals. Hence, subtracting that percentage from 100% and multiplying the result by the total number of COVID-19 deaths generated an estimate of total breakthrough deaths.

The data for the aforementioned 47 jurisdictions indicates that as of March 27, 2022, a minimum of 57,617 COVID-19 breakthrough deaths had occurred in the United States. Note that this includes all deaths among those who had been fully vaccinated, whether or not they were boosted. (Only 10 jurisdictions^7^ reported deaths for boosted and non-boosted individuals separately. Based on this data, there were 10,351 breakthrough deaths among non-boosted people in these 10 jurisdictions, compared to 2047 deaths among people who had been boosted.) The New York Times’ COVID-19 tracker indicates that the total number of COVID-19 deaths in the United States as of March 27, 2022 was 975,515, indicating that at least 57,617/975,515 = 5.91% of all COVID-19 deaths in the United States as of that date were among fully vaccinated individuals.

Hereafter I will define January 1, 2021 as the date when widespread COVID-19 vaccination started to take place in the United States. This seems reasonable partly because it is just a few weeks after the first American got the COVID-19 vaccine (Sandra Lindsay got it on December 14, 2020) (Aubrey 2021), and partly because the KFF (Kates et al. 2021) and numerous other studies (e.g. Watkins et al. 2022, Birhane et al. 2021) have also used this as a starting point for analyzing data related to breakthrough cases.

Furthermore, including only deaths since January 1, 2021, the total number of deaths between that date and March 27, 2022 is 975,515-347,970 = 627,545. Using this denominator, 57,617/627,545 = 9.18% of all deaths in the United States since the start of the vaccine rollout have been among fully vaccinated individuals. For multiple reasons, including the obvious fact that not all U.S. jurisdictions are included in the above data, the true number of breakthrough deaths is certainly higher than the aforementioned minimum.

If we then ignore the fact that even for the 47 jurisdictions for which I could find state-level data, this data does not extend back to the beginning of COVID-19 vaccination in every state, a total of 157,953 of the 970,872 COVID-19 deaths in the United States as of March 27, 2022 had occurred in one of the four states without statewide data. Therefore, 812,919/970,872 = 83.7% of all COVID-19 deaths occurred in the 46 states/DC for which I included data in this paper. So if we assume that the same percentage of breakthrough deaths also occurred in those states, then 57,617 = (83.7%)*(total breakthrough deaths), meaning that the total estimated minimum number of breakthrough deaths in the United States is 57,617/(0.837) = 68,838. Again, this is likely an underestimate of the true number of such deaths in the United States given that it is based on data that (at least for some states) does not cover the entire time period since vaccination against COVID-19 began in the United States. Again considering that breakthrough deaths could only have happened since vaccination started in the United States (defined here as January 1, 2021), the total number of deaths since then is 586,684, as mentioned earlier. Therefore, 68,838/586,684 = 11.7% of COVID-19 deaths since the start of vaccination in the United States are estimated to have been among fully vaccinated individuals.

### Extrapolating the results of Johnson et al. (2022)

Johnson et al. (2022) presented total counts of breakthrough deaths in 25 U.S. jurisdictions: Alabama, Arkansas, California, Colorado, District of Columbia, Florida, Georgia, Idaho, Indiana, Kansas, Louisiana, Massachusetts, Michigan, Minnesota, Nebraska, New Jersey, New Mexico, New York, New York City (New York), Rhode Island, Seattle/King County (Washington), Tennessee, Texas, Utah, and Wisconsin. These data cover the time period April 4, 2021-December 4, 2021. This study reported a total of 22,567 breakthrough deaths in the aforementioned 25 jurisdictions during that time period. Data from the New York Times’ national COVID-19 tracker^8^ and (because the NYT dataset for individual US counties is too big of a file to download or edit effectively) the King County government website indicates that the total number of deaths in all 25 jurisdictions during the April 4-December 4, 2021 time period was 142,798. Thus, a total of 22,567/142,798 = 15.8% of all COVID-19 deaths in this time period and in these jurisdictions were breakthrough deaths. The NYT national dataset indicates that the total number of COVID-19 deaths in the entire United States during this time period was 232,561. So if we assume that 15.8% of all deaths nationally were breakthrough deaths, as was the case with the 25 jurisdictions included in the Johnson et al. (2022) study, 232,561*(15.8%) = 36,745 breakthrough deaths happened in the entire United States during the April 4-December 4, 2021 time period. Furthermore, the total number of COVID-19 deaths in the United States from January 1, 2021 to March 27, 2022 (from the start of vaccination in the United States to the end of the time period analyzed here), as mentioned earlier, is 627,545. So applying the same 15.8% estimate to the national total of deaths during this time period yields 627,545*(15.8%) = 99,152 total breakthrough deaths in the United States.

## Discussion

The data included here from state COVID-19 dashboards and other official sources of data indicates that the total number of COVID-19 breakthrough deaths in the United States as of March 27, 2022 likely exceeded 60,000. Specifically, the minimum number of breakthrough deaths as of that date was 57,617, but given the number of COVID-19 deaths that happened in jurisdictions and/or time periods for which this data is not explicitly reported, this is likely significantly lower than the true number of breakthrough deaths. This minimum number is based on state-level reporting of breakthrough deaths in publicly available data sources such as online COVID-19 dashboards. A study covering most of the year 2021 in many (but not all) U.S. jurisdictions (Johnson et al. 2022) identified a noticeably smaller number of breakthrough deaths, but a significantly higher proportion of all COVID-19 deaths that were in fully vaccinated individuals (15.8%) compared to the proportion based on the dashboard-based minimum estimate (7.9%). Hence, applying the proportion from Johnson et al. implies that 99,152 total breakthrough deaths in the United States took place up until March 27, 2022.

There have been other estimates of the total number of breakthrough COVID-19 deaths in the United States published previously, largely relying on the same patchwork of state dashboards as does the present paper. For example, Mitropoulos (2021) estimated more than 16,700 breakthrough deaths in the United States from April 1, 2021 to November 30, 2021, based on data from 27 U.S. states. Kamp & Evans (2021) similarly published an article on November 21, 2021, reporting a total number of over 20,000 breakthrough deaths across the United States up to that point. It is unsurprising that the total of 57,617 deaths reported here (which is certainly an underestimate due to the multiple states excluded from this data) is significantly larger than those two previous estimates, both because this paper includes more recent data and because it includes data from more U.S. states (at least more states than Mitropoulos; I do not know how many states were used in Kamp & Evans because they do not specify that information) compared to the other two articles.

## Data Availability

I have a Google Sheets spreadsheet including the URLs of the sources for the data in this paper. I can send it to someone if they ask for it.

## Footnotes

2 https://coronavirus.health.ny.gov/covid-19-breakthrough-data

3 https://coronavirus.health.ny.gov/vaccination-progress-date

4 https://www.dshs.texas.gov/immunize/covid19/data/vaccination-status/

5 https://tabexternal.dshs.texas.gov/t/THD/views/COVID-19VaccineinTexasDashboard/Summary?:origin=card_share_link&:embed=y&:isGuestRedirectFromVizportal=y

6 https://web.archive.org/web/20211202204120/https://www.dshs.texas.gov/immunize/covid19/data/Cases-and-Deaths-by-Vaccination-Status-11082021.pdf

7 District of Columbia, Delaware, North Dakota, Mississippi, Utah, New Mexico, Oregon, Connecticut, New Jersey, and California

8 https://github.com/nytimes/covid-19-data/blob/master/us.csv

